# Association of rare variants in *ARSA* with Parkinson’s disease

**DOI:** 10.1101/2023.03.08.23286773

**Authors:** Konstantin Senkevich, Mariia Beletskaia, Aliza Dworkind, Eric Yu, Jamil Ahmad, Jennifer A. Ruskey, Farnaz Asayesh, Dan Spiegelman, Stanley Fahn, Cheryl Waters, Oury Monchi, Yves Dauvilliers, Nicolas Dupré, Lior Greenbaum, Sharon Hassin-Baer, Ilya Nagornov, Alexandr Tyurin, Irina Miliukhina, Alla Timofeeva, Anton Emelyanov, Ekaterina Zakharova, Roy N. Alcalay, Sofya Pchelina, Ziv Gan-Or

## Abstract

**Background:** Several lysosomal genes are associated with Parkinson’s disease (PD), yet the association between PD and *ARSA*, which encodes for the enzyme arylsulfatase A, remains controversial.

**Objectives:** To evaluate the association between rare *ARSA* variants and PD.

**Methods:** To study possible association of rare variants (minor allele frequency<0.01) in *ARSA* with PD, we performed burden analyses in six independent cohorts with a total of 5,801 PD patients and 20,475 controls, using optimized sequence Kernel association test (SKAT-O), followed by a meta-analysis.

**Results:** We found evidence for an association between functional *ARSA* variants and PD in four independent cohorts (P≤0.05 in each) and in the meta-analysis (P=0.042). We also found an association between loss-of-function variants and PD in the UKBB cohort (P=0.005) and in the meta-analysis (P=0.049). However, despite replicating in four independent cohorts, these results should be interpreted with caution as no association survived correction for multiple comparisons. Additionally, we describe two families with potential co-segregation of the *ARSA* variant p.E384K and PD.

**Conclusions:** Rare functional and loss-of-function *ARSA* variants may be associated with PD. Further replication in large case-control cohorts and in familial studies is required to confirm these associations.

## Introduction

Lysosomal genes play a prominent role in the pathogenesis of Parkinson’s disease (PD).^1^ Variants in *GBA1* are amongst the most important risk factors of PD,^2^ and mutations in other lysosomal storage disorder genes have also been associated with PD (e.g. *ASAH1, GALC, SMPD1*).^3-7^ Homozygous or compound heterozygous mutations in *ARSA* may lead to the autosomal recessive lysosomal storage disorder metachromatic leukodystrophy (MLD).^8^ Located on chromosome 22q13.33, the *ARSA* gene encodes arylsulfatase A, which hydrolyzes sulfatides to galactosylceramide and sulfate^8^ (Figure 1). Consequently, hydrolysis of galactosylceramide occurs by the lysosomal enzyme galactosylceramidase, encoded by *GALC*, which is nominated as a PD gene by genome-wide association studies and targeted analyses.^6, 7, 9^

**Figure 1.**
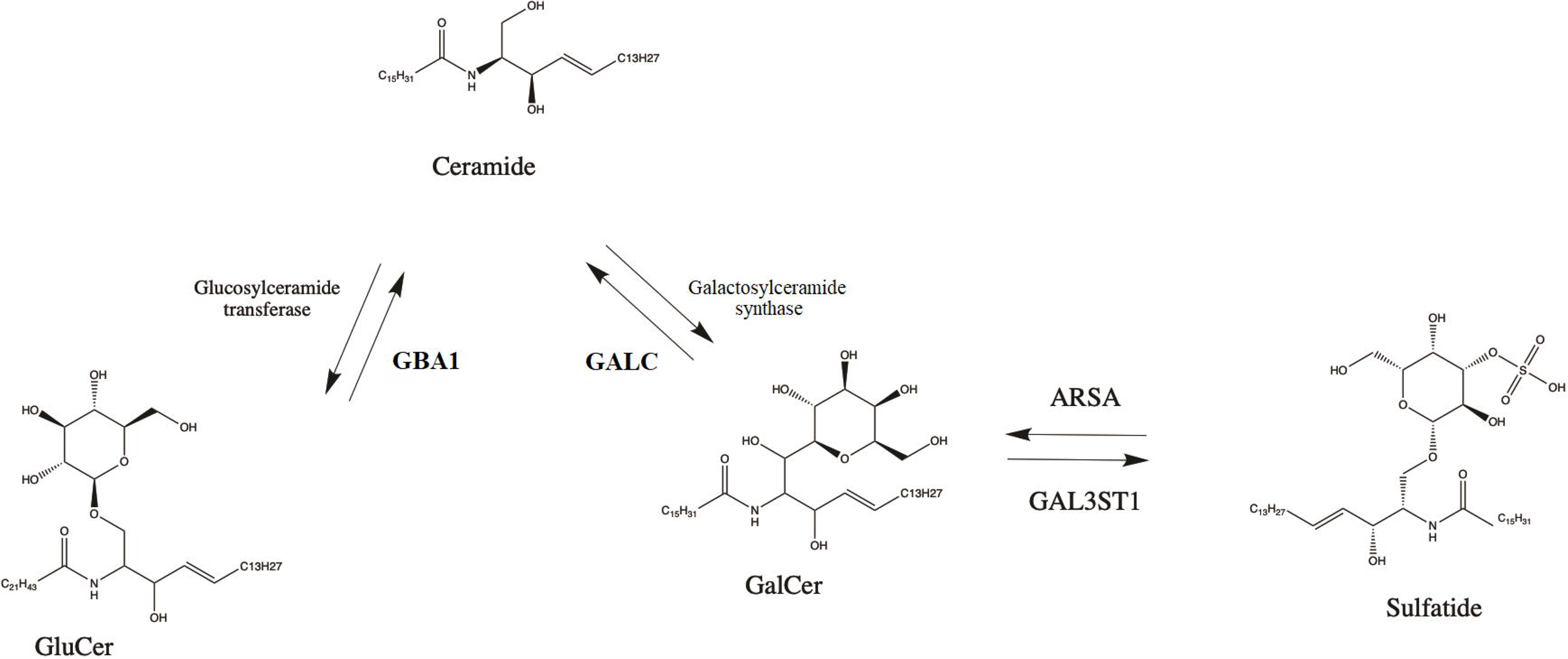
The role of ARSA and GBA1 in sphingolipid metabolism. GluCer- glucosylceramide; GalCer- galactosyleramide; ARSA- arylsulfatase A; GALC- galactosylceramidase; GBA1- galactosylceramidase

The genetic association between *ARSA* variants and PD remains controversial.^10-14^ Co-segregation of pathogenic *ARSA* variant was reported in one family with two PD patients, and two studies suggested potential association between rare *ARSA* loss-of-function variants and PD.^10, 12^ In the current study, we aimed to evaluate the association between rare *ARSA* variants and PD in six cohorts of 5,801 PD patients and 20,475 controls and in two families with MLD and PD.

## Methods

### Population

The study population included a total of 5,801 PD patients and 20,475 controls from six cohorts (detailed in Supplementary Table 1). Four cohorts have been collected and sequenced at McGill University: McGill (Quebec, Canada and Montpellier, France), Columbia University (the SPOT study, New York, NY), Sheba Medical Center (Israel) and Pavlov First State Medical university and Institute of Human Brain (Pavlov and Human Brain cohort; Saint-Petersburg, Russia). Additionally, we analyzed data from the UK Biobank (UKBB) and Accelerating Medicines Partnership – Parkinson Disease (AMP-PD) initiatives. The McGill university cohort was recruited in Québec, Canada (partially through the Quebec Parkinson Network, QPN)^15^ and in France. The Columbia cohort was collected in NY and is of mixed ancestry (European, Ashkenazi Jews [AJ] and a minority of Hispanics and Blacks, described in detail previously)^16^. The Sheba cohort, recruited in Israel, includes only participants with full AJ ancestry (by report). Pavlov and Human Brain cohort, recruited in Russia, consist predominantly of patients of European ancestry. All PD patients in these cohorts were diagnosed by movement disorder specialists according to the UK brain bank criteria^17^ or the MDS clinical diagnostic criteria.^18^ The Accelerating Medicines Partnership – Parkinson Disease (AMP-PD, 2.5 release) initiative cohorts were accessed using the Terra platform (https://amp-pd.org/; AMP-PD cohorts detailed in Acknowledgments). The UKBB cohort was accessed using Neurohub (https://www.mcgill.ca/hbhl/neurohub).

We contacted 21 families with MLD (homozygous or compound heterozygous carriers of pathogenic *ARSA* variants) or their representatives through Russian Society of Rare (Orphan) Diseases and sent them out questionnaire to detect family history of PD. We analyzed *ARSA* mutations using sanger sequencing in two selected families with positive PD history to attempt detection of co-segregation of pathogenic variants within PD patients.

All participants signed informed consent forms before entering the studies and study protocols were approved by the institutional review boards.

### Targeted next generation sequencing

The *ARSA* gene was sequenced in the four cohorts collected at McGill University with targeted next generation sequencing by molecular inversion probes (MIPs) as previously described.^19^ All MIPs that were used to sequence *ARSA* are provided (Supplementary Table 2) and the full protocol is available at https://github.com/gan-orlab/MIP_protocol. The library was sequenced using Illumina NovaSeq 6000 SP PE100 platform at the Genome Quebec Innovation Centre. Alignment was performed with Burrows-Wheeler Aligner (hg19)^20^ and Genome Analysis Toolkit (GATK, v3.8) was used for post-alignment quality control and variant calling.^21^ We performed quality control by filtering out variants and samples with reduced quality, using the PLINK software v1.9. SNPs were excluded from analysis if missingness was more than 10%. Variants with a minor allele frequency (MAF) less than 1% and with a minimum quality score (GQ) of 30 were included in the analyses and analyzed at minimal depths of coverage 30x.

### Data quality control and analysis in AMP-PD and UKBB

Quality control procedures of whole genome sequencing for AMP-PD cohorts were performed on individual and variant levels as described by AMP-PD (https://amp-pd.org/whole-genome-data and detailed elsewhere).^22^ Quality control of UKBB whole exome sequencing data was performed using Genome Analysis Toolkit (GATK, v3.8) with minimum depth of coverage 10x and GQ 20 as described previously^23^ and we removed all multi-allelic sites.

Alignment of AMP-PD and UKBB data was performed using the human reference genome (hg38) and coordinates for the *ARSA* gene extraction were chr22:50,622,754-50,628,152. We performed additional filtration procedures using the UKBB and AMP-PD cohorts to exclude non-European individuals (UKB field 21000) and filtered by relatedness to remove any first and second-degree relatives.

#### Annotations and statistical analysis

To functionally annotate genetic variants in all cohorts, we utilized ANNOVAR.^24^ Data on variant pathogenicity were predicted using Combined Annotation Dependent Depletion (CADD) score and Varsome.^25, 26^ To analyze rare variants (MAF<0.01), an optimized sequence Kernel association test (SKAT-O, R package) was performed.^27^ We separately analyzed the burden of all rare, nonsynonymous and functional variants (nonsynonymous, stop/frameshift and splicing) and loss-of-function variants. Lastly, we analyzed variants with a Combined Annotation Dependent Depletion (CADD) score of ≥ 20, representing the top 1% of potentially deleterious variants. For each of the analyses, we performed a meta-analysis between the cohorts using metaSKAT package,^28^ adjusting for sex, age and ethnicity. We applied false discovery rate (FDR) correction to all p-values. All the code used in the current study is available at https://github.com/gan-orlab/ARSA

## Results

### Rare functional and loss-of-function *ARSA* variants are associated with Parkinson’s disease

The average coverage across all four cohorts sequenced at McGill was >714X with >98% of the nucleotides covered at >30x (detailed in Supplementary Table 3). We identified a total of 96 rare variants across all cohorts sequenced at McGill (Supplementary Table 4) and 113 rare variants in AMP-PD and UKBB cohorts (Supplementary Table 5).

Burden analyses, using SKAT-O, demonstrated an association of functional variants with PD in four out of six cohorts (McGill, P=0.023, Columbia, P=0.037, Pavlov, P=0.022 and UKBB, P=0.009) and in the meta-analysis (P=0.042; Table 1; Supplementary Table 6). We also found an association between rare loss-of-function variants in the UKBB cohort (P=0.005) and in the meta-analysis (P=0.049). However, these results should be interpreted with caution as only a single loss-of-function variant was reported in the Columbia cohort, two in Pavlov and Human brain cohort, three variants in UKBB and two in AMP-PD (Supplementary Tables 4-5) and none of the associations survived FDR correction (Supplementary Table 6).

**Table 1.** Burden analysis of rare *ARSA* variants.

| Cohort | N cases | N controls | All rare variants, P | All non-synonymous variants, P | Functional variants, P | Loss of function, P | CADD > 20, P |
| --- | --- | --- | --- | --- | --- | --- | --- |
| Columbia cohort | 917 | 486 | 0.005 | 0.060 | 0.037 | 0.313 | 0.009 |
| Sheba cohort | 683 | 553 | 0.195 | 0.745 | 0.095 | - | 0.664 |
| McGill cohort | 761 | 549 | 0.011 | 0.032 | 0.023 | - | 0.081 |
| Pavlov and Human brain cohort | 497 | 401 | 0.019 | 0.106 | 0.022 | 0.467 | 0.082 |
| UKBB | 602 | 15,000 | 0.009 | 0.686 | 0.009 | 0.005 | 0.539 |
| AMP-PD | 2,341 | 3,486 | 0.820 | 0.673 | 0.602 | 0.107 | 0.705 |
| Meta-analysis of all cohorts | 5,801 | 20,475 | 0.826 | 0.420 | 0.042 | 0.049 | 0.431 |

N, number; P, p value; UKBB, UK biobank; AMP-PD, Accelerating Medicines Partnership – Parkinson Disease; CADD, Combined Annotation Dependent Depletion score.
p-value presented without FDR adjustment, as no p-values survived after correction.

We found associations between all rare variants and PD in the McGill cohort (P=0.011), Columbia cohort (P=0.005), Pavlov and Human brain institute (P=0.019) and in the UKBB cohort (P=0.009). However, there was no association in the meta-analysis (Table 1; Supplementary Table 4). Variants with CADD scores ≥20 were associated with PD in the Columbia cohort (P=0.009), whereas no association was found in the other cohorts and in the meta-analysis. Similarly, all rare nonsynonymous variants in *ARSA* were associated with PD in the McGill cohort (P=0.032) but not in the other cohorts. We did not find the p.L300S *ARSA* variant, which was previously reported as pathogenic in PD,^29^ yet we found the likely pathogenic (based on Varsome annotation) p.L300V variant in two cases and one control in our analysis (Supplementary Tables 5-6).

### Evidence for association of the rare *ARSA* p.E384K in two families among Parkinson’s disease patients

We describe here two families with history of MLD and PD. In the first family (Figure 2A), the proband is a patient with MLD with compound heterozygous nonsynonymous variants, p.Q155H and p.E384K. The maternal grandmother of the proband (Figure 2A, II-4), who is a carrier of p.E384K, has PD. The patient had early PD onset (<50 years). Other healthy relatives in this maternal generation (II) were wildtype for this variant. In the second family, the proband is a MLD patient who has compound heterozygous mutations, c.1107+1G>A and p.E384K. There were five PD patients in this family from both the paternal and maternal sides (Figure 2B). On the paternal side, there were two PD patients, one was deceased, and one was not a carrier of the pathogenic variant c.1107+1G>A. On the maternal side, there were three PD patients, all deceased. The maternal grandmother was wildtype to this variant. Therefore, the grandfather who was a PD patient was likely a carrier of p.E384K.

**Figure 2.**
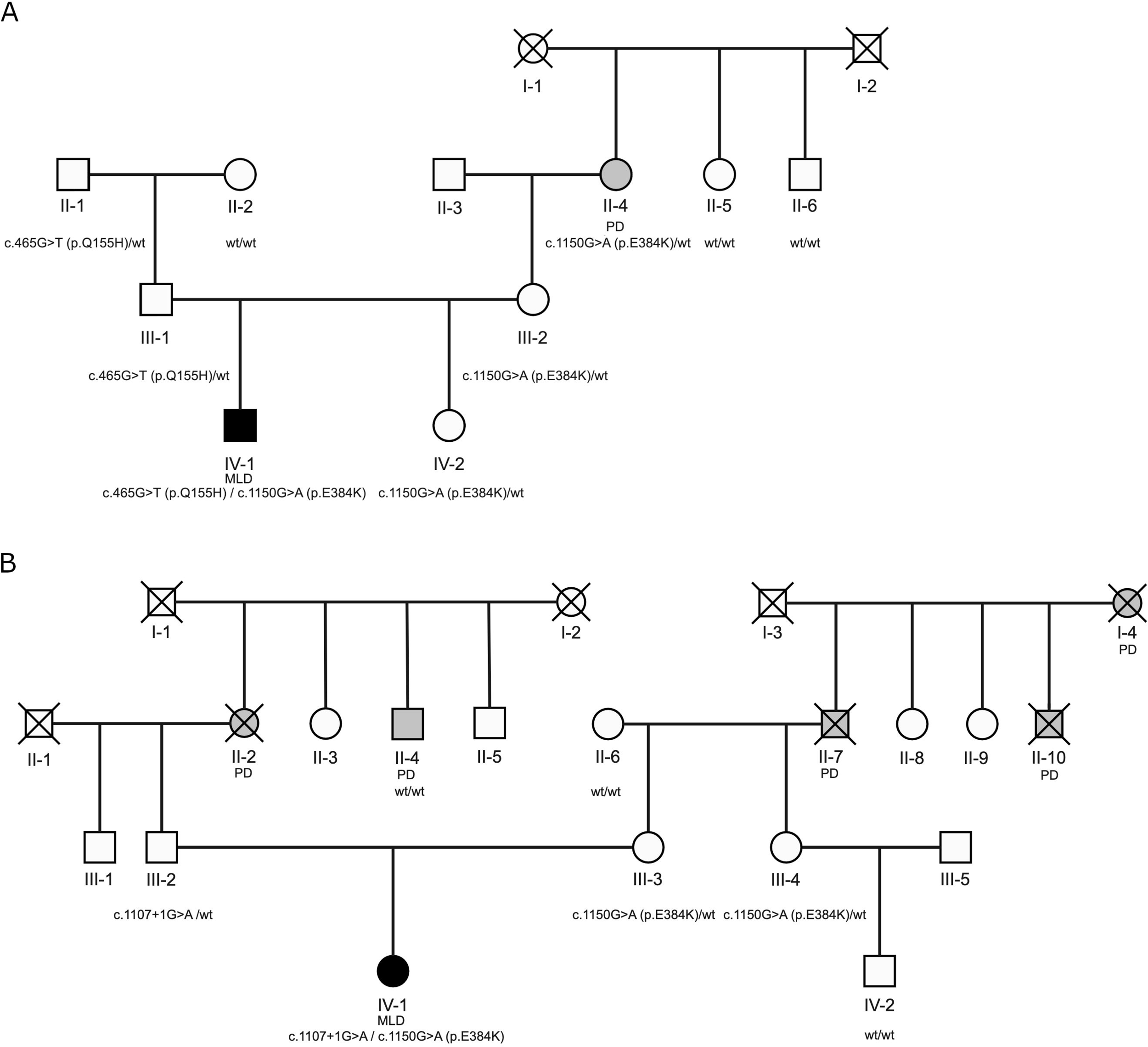
Family trees of two families with Metachromatic leukodystrophy and Parkinson’s disease in history. Square – male; circle – female; open symbol – healthy; grey– Parkinson’s disease; filled black symbol – Metachromatic leukodystrophy; PD – Parkinson’s disease; MLD – Metachromatic leukodystrophy; crossed line – deceased subject; wt – wild-type.

## Discussion

In the current study, we report a possible association between rare functional and loss-of-function *ARSA* variants and PD. In four of our cohorts, we also identified a possible association between all rare and nonsynonymous variants and PD. We also found a potential segregation of a pathogenic variant, p.E384K, with PD in 2 families with family history of PD and MLD, albeit we could not confirm this for all affected family members as some of the family members with PD have passed away. The negative results previously reported for rare *ARSA* variants in PD could be attributed to sample size or ethnicity (Supplementary Table 7).^12-14^ Although the associations described in the present study do not survive correction for multiple comparisons, the fact that there were many nominal associations in independent cohorts may suggest that these associations are real.

A recent large scale burden analysis found an association between rare *ARSA* loss-of-function variants and PD.^10^ While a study from China did not find a statistically significant burden of rare *ARSA* variants in PD,^30^ they reported higher prevalence of loss-of-function variants in late-onset PD (0.25% in PD vs 0% in controls),^30^ which is in line with our results. However, our results should be interpreted with caution as none of our associations survived FDR correction and we only discovered a few carriers of private loss-of-function variants across all six cohorts. A recent study from Japan suggested that the *ARSA* p.L300S mutation was likely pathogenic in PD due to co-segregation within a family with two PD patients.^29^ We did not find this specific variant in our study. However, it is possible that the variant p.E384K could be associated with PD based on the data we gathered from two families with MLD and PD.

The enzyme encoded by *ARSA*, arylsulfatase A, has an important role in the lysosomal ceramide metabolism pathway. Galactosylceramide is hydrolyzed from sulfatides by arylsulfatase A, which is then further hydrolyzed to ceramide by galactosylceramidase,^31^ encoded by the putative PD gene *GALC*.^7^ Another PD gene, *GBA1*,^1, 32^ also plays an important role in ceramide metabolism, by hydrolyzing glucosylceramide to ceramide (Figure 1). *ARSA* is also important for myelin metabolism.^33^ Several studies suggested a link between *ARSA* and alpha-synuclein accumulation. Alpha-synuclein depositions were found in glial cells and microglia of MLD patient,^34^ and in *ARSA* knockout cells, the authors reported increased alpha-synuclein accumulation, secretion and propagation.^11^ The activity of ARSA was reported to be low in the subset of patients with parkinsonism.^35^ Moreover, plasma ARSA level was reported to be higher in early PD as compared to controls or late PD, suggesting possible compensatory mechanism.^36^ Reduced level of sulfatides, substrate of ARSA, was reported in frontal cortex of PD patients.^37^ Therefore, there is biochemical, functional, and genetic evidence for the involvement of *ARSA* in neurodegeneration and potentially PD, further emphasizing the importance of the lysosomal ceramide metabolism pathway in PD (Figure 2). The link between *ARSA* and PD is not as strong as between *GBA1* and PD and only evident in large scale burden analysis (Supplementary Table 7). Potentially, it could be due to rarity of *ARSA* variants that associated with PD and could depend on the ethnicity.

Our study has several limitations. In some of our cohorts, patients and controls were not matched for sex and age, which was therefore adjusted in the statistical analysis. Quality control procedures were performed independently for targeted sequencing, whole exome and whole genome sequencing data using different thresholds for depth of coverage and quality control. This could potentially lead to discrepancy in enrichment in variants between different cohorts. Another limitation of our study is the inclusion of mainly individuals of European ancestry.

To conclude, rare functional and loss of function *ARSA* variants may be associated with PD, yet the results here cannot be considered as conclusive. Further replications in other cohorts are required to confirm our findings along with additional functional studies to understand the potential mechanism.

## Data Availability

All data produced in the present work are contained in the manuscript

https://github.com/gan-orlab/ARSA

## Acknowledgement

We would like to thank the participants in the different cohorts for contributing to this study. This research used the NeuroHub infrastructure and was undertaken thanks in part to funding from the Canada First Research Excellence Fund, awarded through the Healthy Brains, Healthy Lives initiative at McGill University, Calcul Québec and Compute Canada. UK Biobank Resources were accessed under application number 45551. Data used in the preparation of this article were obtained from the AMP PD Knowledge Platform. For up-to-date information on the study, visit https://www.amp-pd.org. AMP PD – a public-private partnership – is managed by the FNIH and funded by Celgene, GSK, the Michael J. Fox Foundation for Parkinson’s Research, the National Institute of Neurological Disorders and Stroke, Pfizer, Sanofi, and Verily. Genetic data used in preparation of this article were obtained from the Fox Investigation for New Discovery of Biomarkers (BioFIND), the Harvard Biomarker Study (HBS), the Parkinson’s Progression Markers Initiative (PPMI), the Parkinson’s Disease Biomarkers Program (PDBP), the International LBD Genomics Consortium (iLBDGC), and the STEADY-PD III Investigators. BioFIND is sponsored by The Michael J. Fox Foundation for Parkinson’s Research (MJFF) with support from the National Institute for Neurological Disorders and Stroke (NINDS). The BioFIND Investigators have not participated in reviewing the data analysis or content of the manuscript. For up-to-date information on the study, visit michaeljfox.org/news/biofind. The HBS is a collaboration of HBS investigators [full list of HBS investigator found at https://www.bwhparkinsoncenter.org/biobank/ and funded through philanthropy and NIH and Non-NIH funding sources. The HBS Investigators have not participated in reviewing the data analysis or content of the manuscript. PPMI – a public-private partnership – is funded by the Michael J. Fox Foundation for Parkinson’s Research and funding partners, including [list the full names of all of the PPMI funding partners found at www.ppmi-info.org/fundingpartners]. The PPMI Investigators have not participated in reviewing the data analysis or content of the manuscript. For up-to-date information on the study, visit www.ppmi-info.org. PDBP consortium is supported by the NINDS at the National Institutes of Health. A full list of PDBP investigators can be found at https://pdbp.ninds.nih.gov/policy. The PDBP investigators have not participated in reviewing the data analysis or content of the manuscript. Genome Sequencing in Lewy Body Dementia and Neurologically Healthy Controls: A Resource for the Research Community.” was generated by the iLBDGC, under the co-directorship by Dr. Bryan J. Traynor and Dr. Sonja W. Scholz from the Intramural Research Program of the U.S. National Institutes of Health. The iLBDGC Investigators have not participated in reviewing the data analysis or content of the manuscript. For a complete list of contributors, please see: bioRxiv 2020.07.06.185066; doi: https://doi.org/10.1101/2020.07.06.185066. STEADY□PD III is a 36□month, Phase 3, parallel group, placebo□controlled study of the efficacy of isradipine 10 mg daily in 336 participants with early Parkinson’s Disease that was funded by the NINDS and supported by The Michael J Fox Foundation for Parkinson’s Research and the Parkinson’s Study Group. The STEADY-PD III Investigators have not participated in reviewing the data analysis or content of the manuscript. The full list of STEADY PD III investigators can be found at: https://clinicaltrials.gov/ct2/show/NCT02168842. Pavlov and Human Brain cohort is supported by the Ministry of Health of the Russian Federation (Project No 123030200067-6). We would like to thank All-Russian Society of Rare (Orphan) Diseases for providing assistance and information. ZGO is supported by the Fonds de recherche du Québec - Santé (FRQS) Chercheurs-boursiers award, in collaboration with Parkinson Quebec, and is a William Dawson Scholar. The access to part of the participants for this research has been made possible thanks to the Quebec Parkinson’s Network (http://rpq-qpn.ca/en/). KS is supported by a post-doctoral fellowship from the Canada First Research Excellence Fund (CFREF), awarded to McGill University for the Healthy Brains for Healthy Lives initiative (HBHL) and FRQS post-doctoral fellowship.

## Authors’ Roles

1. Research project: **A**. Conception, **B**. Organization, **C**. Execution
2. Statistical Analysis: **A**. Design, **B**. Execution, **C**. Review and critique
3. Manuscript Preparation: **A**. Writing of the first draft, **B**. Review and critique

KS: 1A, 1B, 1C, 2A, 2B, 3A

MB: 1C, 2B, 3B

AD: 1C, 2B, 2C, 3A

EY: 1C, 2C, 3B

JA: 1C, 2C, 3B

JAR: 1C, 2C, 3B

FA: 1C, 2C, 3B

DS: 1C, 2C, 3B

SF: 1C, 2C, 3B

CW: 1C, 2C, 3B

OM: 1C, 2C, 3B

YD: 1C, 2C, 3B

ND: 1C, 2C, 3B

LG: 1C, 2C, 3B

SHB: 1C, 2C, 3B

IN: 1C, 2C, 3B

AT: 1C, 2C, 3B

IM: 1C, 2C, 3B

AT: 1C, 2C, 3B

AE: 1C, 2C, 3B

EZ: 1B, 1C, 2C, 3B

RNA: 1A, 1C, 2C, 3B

SP: 1B, 1C, 2C, 3B

ZGO: 1A, 1B, 2A, 2C, 3A, 3B

## Financial Disclosures of all authors (for the preceding 12 months)

ZGO received consultancy fees from Lysosomal Therapeutics Inc. (LTI), Idorsia, Prevail Therapeutics, Inceptions Sciences (now Ventus), Ono Therapeutics, Bial Biotech, Bial, Handl Therapeutics, UCB, Capsida, Denali Lighthouse, Guidepoint and Deerfield. Other authors have nothing to disclose.

## Supplementary Tables

Supplementary Table 1 Study population

Supplementary Table 2 Detailed information on the *ARSA* molecular inversion probes

Supplementary Table 3 Coverage details for *ARSA*

Supplementary Table 4 Rare *ARSA* variants for cohorts sequenced at McGill

Supplementary Table 5 Rare *ARSA* variants for UKBB and AMP-PD cohorts

Supplementary Table 6 Burden analysis

Supplementary Table 7 Previous rare variants analysis of *ARSA* in PD

## Notes

**Relevant conflicts of interest/financial disclosures:** RNA research is funded by the Michael J. Fox Foundation, the Silverstein Foundation and the Parkinson’s Foundation. He received consultation fees from Gain Therapeutics, Takeda and Genzyme/Sanofi. ZGO received consultancy fees from Lysosomal Therapeutics Inc. (LTI), Idorsia, Prevail Therapeutics, Inceptions Sciences (now Ventus), Ono Therapeutics, Bial Biotech, Bial, Handl Therapeutics, UCB, Capsida, Denali Lighthouse, Guidepoint and Deerfield. The rest of the authors have nothing to report.

**Funding:** This study was financially supported by grants from the Michael J. Fox Foundation, the Canadian Consortium on Neurodegeneration in Aging (CCNA), the Canada First Research Excellence Fund (CFREF), awarded to McGill University for the Healthy Brains for Healthy Lives initiative (HBHL), and Parkinson Canada. The Columbia University cohort is supported by the Parkinson’s Foundation, the National Institutes of Health (K02NS080915 and UL1 TR000040) and the Brookdale Foundation.

### Competing Interest Statement

RNA research is funded by the Michael J. Fox Foundation, the Silverstein Foundation and the Parkinson’s Foundation. He received consultation fees from Gain Therapeutics, Takeda and Genzyme/Sanofi. ZGO received consultancy fees from Lysosomal Therapeutics Inc. (LTI), Idorsia, Prevail Therapeutics, Inceptions Sciences (now Ventus), Ono Therapeutics, Bial Biotech, Bial, Handl Therapeutics, UCB, Capsida, Denali Lighthouse, Guidepoint and Deerfield. The rest of the authors have nothing to report.

### Funding Statement

This study was financially supported by grants from the Michael J. Fox Foundation, the Canadian Consortium on Neurodegeneration in Aging (CCNA), the Canada First Research Excellence Fund (CFREF), awarded to McGill University for the Healthy Brains for Healthy Lives initiative (HBHL), and Parkinson Canada. The Columbia University cohort is supported by the Parkinson′s Foundation, the National Institutes of Health (K02NS080915 and UL1 TR000040) and the Brookdale Foundation.

### Author Declarations

The study was approved by McGill university Institutional Review Board (IRB), approval A11-M60-21A.

